# Towards individualized Medicine in Stroke – the TiMeS project: protocol of longitudinal, multi-modal, multi-domain study in stroke

**DOI:** 10.1101/2022.05.18.22274612

**Authors:** L Fleury, PJ Koch, MJ Wessel, C Bonvin, D San Millan, C Constantin, P Vuadens, J Adolphsen, AG Cadic-Melchior, J Brügger, E Beanato, M Ceroni, P Menoud, D de Leon Rodriguez, V Zufferey, N Meyer, P Egger, S Harquel, T Popa, E Raffin, G Girard, JP Thiran, C Vaney, V Alvarez, J-L Turlan, A Mühl, B Leger, T Morishita, S Micera, O Blanke, D Van de Ville, FC Hummel

**Affiliations:** Defitech Chair for Clinical Neuroengineering, Center for Neuroprosthetics (CNP) and Brain Mind Institute (BMI), École polytechnique fédérale de Lausanne (EPFL), Geneva, Switzerland; Defitech Chair for Clinical Neuroengineering, CNP and BMI, EPFL Valais, Clinique Romande de Réadaptation, Sion, Switzerland Swiss Federal Institute of Technology (EPFL Valais), Sion, Switzerland; Department of Neurology, University of Lübeck, Lübeck, Germany; Department of Neurology, Julius-Maximilians-University Wu□rzburg, Wu□rzburg, Germany; Hôpital du Valais, Sion, Switzerland; Clinique Romande de Réadaptation, Sion, Switzerland; Berner Klinik, Crans-Montana, Switzerland; Laboratory of Cognitive Neuroscience, Brain Mind Institute & Center for Neuroprosthetics, Ecole Polytechnique Fédérale de Lausanne (EPFL), Campus Biotech, Geneva, Switzerland; CIBM Center for Biomedical Imaging, Switzerland; Radiology Department, Centre Hospitalier Universitaire Vaudois and University of Lausanne, Lausanne, Switzerland; Signal Processing Laboratory (LTS5), Ecole Polytechnique Fédérale de Lausanne (EPFL), Switzerland; The Biorobotics Institute and Department of Excellence in Robotics & AI, Scuola Superiore Sant’Anna, Pisa, Italy; Bertarelli Foundation Chair in Translational Neuroengineering, Centre for Neuroprosthetics and Institute of Bioengineering, School of Engineering, Ecole Polytechnique Fedeerale de Lausanne (EPFL), Lausanne, Switzerland; Department of Neurology, University of Geneva (UNIGE), Geneva, Switzerland; Medical Image Processing Lab, Institute of Bioengineering, Center for Neuroprosthetics, Ecole Polytechnique Fédérale de Lausanne, Lausanne, VD, Switzerland; Department of Radiology and Medical Informatics, University of Geneva (UNIGE), Geneva, Switzerland; Clinical Neuroscience, Geneva University Hospital, Geneva, Switzerland

**Author notes:** Corresponding author: Friedhelm C. Hummel.

**Keywords:** stroke, precision medicine, transcranial magnetic stimulation, electroencephalography, neuroimaging, biomarkers, recovery, neuropsychology

## Abstract

Despite recent improvements, complete motor recovery occurs in less than 15% of stroke patients. To improve the therapeutic outcomes, there is a strong need to tailor treatments to each individual patient. However, there is a lack of knowledge concerning the precise neuronal mechanisms underlying the degree and course of motor recovery and its individual differences, especially in the view of network properties despite the fact that it became more and more clear that stroke is a network disorder. The TiMeS project is a longitudinal exploratory study aiming at characterizing stroke phenotypes of a large, representative stroke cohort through an extensive, multi-modal and multi-domain evaluation. The ultimate goal of the study is to identify prognostic biomarkers allowing to predict the individual degree and course of motor recovery and its underlying neuronal mechanisms paving the way for novel interventions and treatment stratification for the individual patients. A total of up to 100 patients will be assessed at 4 timepoints over the first year after the stroke: during the first (T1) and third (T2) week, then three (T3) and twelve (T4) months after stroke onset. To assess underlying mechanisms of recovery with a focus on network analyses and brain connectivity, we will apply synergistic state-of-the-art systems neuroscience methods including functional, diffusion, and structural magnetic resonance imaging (MRI), and electrophysiological evaluation based on transcranial magnetic stimulation (TMS) coupled with electroencephalography (EEG) and electromyography (EMG). In addition, an extensive, multi-domain neuropsychological evaluation will be performed at each timepoint, covering all sensorimotor and cognitive domains. This project will significantly add to the understanding of underlying mechanisms of motor recovery with a strong focus on the interactions between the motor and other cognitive domains and multimodal network analyses. The population-based, multi-dimensional dataset will serve as a basis to develop biomarkers to predict outcome and promote personalized stratification towards individually tailored treatment concepts using neuro-technologies, thus paving the way towards personalized precision medicine approaches in stroke rehabilitation.

## Introduction and rationale

With 80 million survivors in 2016, stroke is the second most common cause of acquired disabilities in the world (1,2). This number is still increasing due to the population growth and ageing (3). Better acute stroke management results in an improved stroke survival, but implies a higher prevalence of chronic stroke (2). Yet, complete motor recovery still occurs in less than 15% of patients (4). Moreover, although motor deficits are the most debilitating and investigated (5–7), patients also show consistent long-lasting cognitive deficits (8,9), with a relevant proportion of patients having multiple domains affected. These long-term impairing behavioral deficits have a strong impact on patients’ reintegration, on patients and their relatives’ daily life, but also on socioeconomics and health care systems (10,11). Therefore, the call for effective strategies of neurorehabilitation in order to maximize the rate of recovery is recognized as a priority to substantially reduce the burden of stroke survivors (2,12). However, the heterogeneity in stroke outcome and in individual recovery potential is an important challenge to address, in order to provide optimal rehabilitative therapies. A crucial aspect to take up this challenge is to deepen our understanding of individual courses of recovery and the underlying neuronal mechanisms through the identification of associated biomarkers (13).

On the behavioral level, stroke is known to yield multiple deficits. The most reported and debilitating ones are the motor impairments, present in 50% to 80% of stroke survivors (7). In particular, damages to the upper extremity function are common and significantly impact the patients’ capacity to retrieve independence, as well as to reintegrate to professional life (14,15). Besides motor deficits, cognitive impairment is common in stroke survivors although initially less obvious: half of stroke survivors report difficulties in at least one cognitive domain, but this area is much less studied than the motor domain (8,16). Cognitive impairment could be found in multiple domains most frequently in, e.g., executive functions, attentional functions or memory. Such deficits are significantly persistent after one to several years after the stroke (8,17). Cognitive deficits also represent an obstacle for patients to go back into a normal daily life (10,18,19). Furthermore, these dysfunctions might strongly impact, slow or even prevent proper motor recovery and response to treatment (20). For example, it is known that executive functions, such as information processing and motor planning are essential in the processes of motor (re)learning (21), which is crucial in motor rehabilitation following stroke. However, despite few investigations of the relationships between these domains (e.g. 17,22), research mainly focused so far on deficits in only one domain, e.g. motor (23), language (24) or attention (25) and neglected largely the interaction between them. Thus, there is a strong lack of knowledge about how deficits in different domains depend on and influence each other in regard of impairment, residual functions and the process of regaining lost functions after a stroke.

Recovery is often incomplete among stroke survivors, and the potential of restoring lost functions is crucially highly heterogeneous between patients (26,27). For example, spontaneous natural recovery in motor domain occurs in roughly 2/3 of patients (13) who recover about ∼70% in average of their maximum recovery potential given their initial impairment (28). In contrast, roughly 1/3 of patients presents altered or insufficient intrinsic plasticity after stroke leading to a poor natural recovery (13). Such heterogeneity has also been reported in other cognitive deficits e.g., neglect and aphasia (29). In addition, stroke survivors act highly heterogeneous in the view of the response towards specific treatment strategies, resulting in the distinction between responders and non-responders (30–32). For instance, patients with cortical lesions specifically demonstrated low responsiveness to repetitive Transcranial Magnetic Stimulation (TMS) protocols (33). Therefore, a key challenging aspect for enhancing neuro-rehabilitation efficacy might be to shed light on the heterogeneity of stroke patients and leverage this information to determine and predict the degree of impairment and potential for individual functional recovery (34,35). This heterogeneity in stroke ranges from brain reorganization to behavioral outcomes and needs to be accounted for when planning rehabilitation strategies (32,34).

The identification of specific individual patterns of recovery through a multi-domain perspective during the first weeks/months post-stroke, and crucially the uncovering of the underlying brain reorganization mechanisms would be a massive step towards the optimization of treatment strategies for each patient. However, there is a lack of understanding concerning the detailed neuronal mechanisms following a stroke lesion and during the course of recovery. Accumulating evidence suggests that stroke is not a focal disorder, but a network disorder (36,37). In addition to local brain tissue damage, stroke also impacts the functioning of connected areas (close or remote from the lesion) as a result of alterations in brain networks (38). In addition, functional reorganization associated with recovery is also not restricted to a focal area. For instance, cortical plasticity associated with motor recovery is not restricted to the primary motor cortex (M1), but rather embraces the complete motor network, including primary and secondary motor cortical areas in both hemispheres, subcortical areas like the basal ganglia and the cerebellum (35, 39, 40). Factors such as lesion size and location (e.g., 41,42), as well as structural and functional prerequisites and dynamics (43) might relevantly influence recovery-associated plasticity processes in the brain leading to heterogeneous, widespread and time-dependent changes of brain reorganization and connectivity between patients. To improve rehabilitative strategies, it is therefore crucial to take this heterogeneity into account and understand how it relates to the pattern of network reorganization and the range of behavioral outcomes following a stroke.

On the basis of this reasoning, there is a strong need for an exact phenotyping of patients that would consider stroke heterogeneity in order to predict outcome and course of recovery and to further improve stroke recovery and treatment outcomes. Such challenge requires to gain a detailed and fundamental knowledge about the precise neuronal mechanisms associated with behavioral recovery, with a particular emphasis on brain networks changes. In addition, is essential to investigate the different domains impacted by the stroke instead of focusing on one behavioral outcome. As network and behavioral alterations following stroke are dynamic and not linear, a longitudinal investigation is of great importance. Such phenotyping will allow to distinguish distinct profiles of patients with associated dynamics of brain reorganization over the course of recovery. Enhancing the fundamental knowledge of stroke diversity through a multimodal and multidomain approach would serve as a basis to pave the way for personalized precision medicine in the field of stroke recovery to achieve maximal treatment effects.

To take up this challenge, the TiMeS project aims at characterizing in details phenotypes of stroke patients allowing to determine the individual course and degree of recovery following stroke and to identify relevant biomarkers associated with recovery. To that purpose, the goal is to collect a large multidimensional dataset that would be representative for the stroke population. Measurements will come from synergistic state-of-the-art systems neuroscience methods including magnetic resonance imaging (MRI), transcranial magnetic stimulation (TMS) coupled with electroencephalography (EEG), in a longitudinal assessment from acute to chronic stage during the first year after the stroke. As stroke is not a focal disorder, subsequent analyses will focus on networks properties within the whole brain and their changes over time, in combination with stroke behavioral outcomes. To provide detailed knowledge about the behavioral patterns and relationships between domains, the procedure will contain an extensive evaluation of behavioral outcomes in multiple domains, including a multi-cognitive assessment. The multidimensional dataset acquired through this research will enable to assess for the first time the complex interactions of structural and functional brain connectivity parameters within certain domain-specific networks as well as within the whole brain, and to associate them with stroke behavioral outcomes and functional recovery.

## Methods

### Study design

The present project is an on-going longitudinal observational study. We follow-up a total of up to 100 stroke patients at four timepoints over one year after the ictal event (T1: 1^st^ week, T2: three weeks, T3: three months, T4: twelve months) from the acute to the chronic phase of recovery. At each timepoint, we investigate the neural correlates of recovery and the underlying plasticity through a multi-modal and multi-domain set of evaluations including structural, diffusion, and functional neuroimaging (MRI), electrophysiology (resting-state EEG, and TMS coupled with EEG) and an extensive battery of tests assessing the multi-domain functional and behavioral outcomes of the patients.

### Objectives

The main goal of the study is to assess the inter-individual variance and different phenotypes of patients after a stroke (ischemic or hemorrhagic). The main goal is divided into three related objectives: 1) to evaluate the neuro-imaging and neurophysiological factors that determine the course and degree of recovery with a focus on structural and functional connectomics, 2) to determine the interactions between multiple cognitive, visual, sensory, and motor functions and their impact on impairment, residual functions and recovery, and 3) to associate the changes in connectomics parameters with the clinical-behavioral outcomes.

To complete these objectives, we apply a multimodal assessment of neuro-imaging and neurophysiological parameters to leverage the advantages of each methods and account for their specific limitations to achieve a very detailed picture, especially in the view of the importance of network analyses. In addition, we use an extensive battery of behavioral tests to acquire detailed information concerning the patients’ motor and cognitive profiles as well as their dynamics. The overall goal of this research will be to integrate and combine the multimodal data (i.e. neuroimaging, electrophysiology, and behavioral) together to obtain detailed and complete phenotypes of stroke patients.

### Primary outcome

As upper extremity function and impairment are the main reason for long-term disability and predictors of reintegration in normal life and functional independence after stroke, longitudinal recovery of the upper limb function and its underlying mechanisms are the primary interest of this study. Upper limb motor function includes multiple aspects, fine and gross dexterity, gross motor function, strength, spasticity, etc (44). These aspects are assessed longitudinally using the same set of reliable and validated clinical tests at each timepoint (see *Appendix n°1* for details). We are especially interested in how other cognitive domains and their alterations after a stroke impact on motor recovery.

### Secondary outcomes

Secondary outcomes are specific readouts based on the multi-domain cognitive evaluation and the multi-modal data from system neurosciences techniques, i.e. neuro-imaging and electrophysiological methods.

#### Magnetic Resonance Imaging (MRI)

Structural, diffusion-weighted and resting-state functional MRI are used to obtain individual structural and functional network properties to evaluate lesion-related neuronal alterations as well as their dynamics throughout the recovery phase, i.e. neuronal plasticity, reorganization and degeneration. Analyses will mainly focus on brain network alterations and changes over time through disconnectomics (45) and by applying computational approaches such as graph theory methods (46), e.g the *Rich-Club* approach (47). In addition, integrated analyses of brain structure and function will be emphasized, e.g. by using the Structural Decoupling Index (SDI), a metric that allows to quantify the coupling strength between structure and function (48). MRI methods and sequences are detailed in *Appendix n°2*.

#### Electrophysiological recordings

Functional measurements of the cortical excitability are provided by means of Transcranial Magnetic Stimulation (TMS). We use single pulses delivered to the primary motor cortex (M1) to generate motor evoked potential (MEPs) and to screen for cortico-spinal tract integrity. We also apply paired-pulses to assess the short-interval intracortical inhibition (SICI; 49). This is thought to reflect GABA_A_-mediated inhibition in the motor cortex (50). Electroencephalography (EEG) allows to assess the resting state brain connectivity (51). More importantly, in combination with TMS, EEG is used to assess interregional connectivity in the brain and to characterize the TMS-evoked potential and its evolution during the course of recovery. Therefore, TMS-EEG represents a unique method to study brain dynamics and their changes over time as it allows to record directly and non-invasively various neurophysiological processes across motor and non-motor areas e.g. cortical responsiveness, cortico-cortical interactions, local excitation and inhibition, oscillatory activity etc (see Tremblay et al., 2019 for a recent review). Electrophysiological methods are detailed in *Appendix n°3*.

#### Behavioral outcomes

To assess precisely the motor and cognitive profiles of the patients, an extensive battery of 40 tests is performed at each timepoint by a trained neuropsychologist. The battery covers sensory-motor domains as well as each neuro-cognitive domain as defined in the DSM-V, i.e. executive functions, language, complex attention, learning and memory, social cognition, perceptual-motor domains (53). Multiple questionnaires complete this battery to evaluate additional aspects such as fatigue, mood, functional independence and recovery. See *Appendix* n°1 for details.

### Study organization

#### Ethical considerations

The study was designed and is conducted according to the guidelines of the Declaration of Helsinki. All the procedures were approved by the cantonal ethics committee (Project ID 2018-01355).

#### Eligibility

We look for stroke patients presenting some upper limb motor impairment in the acute stage. In order to get a heterogeneous cohort, we screen patients with first-ever as well as recurrent stroke, either ischemic or hemorrhagic. Detailed inclusion and exclusion criteria are following:

- Inclusion criteria
  - Age > 18 years old
  - First-ever or recurrent stroke
  - Ischemic or hemorrhagic stroke
  - Stroke incident < 7 days at consent
  - Motor impairment in the acute stage, objectified by a clinical assessment
  - Absence of contraindication for NIBS and MRI
- Exclusion criteria
  - Severe neuropsychiatric (e.g. major depression, severe dementia) or medical disease
  - Not able to consent
  - Severe sensory or cognitive impairment or musculoskeletal dysfuntions prohibiting to understand instructions or the perform the experimental tasks
  - Implanted medical electronic devices or ferromagnetic metal implants, which are not MRI and TMS compatible
  - History of seizures
  - Medication that significantly interacts with NIBS being benzodiazepines, tricyclic antidepressant and antipsychotics
  - Pregnancy
  - Regular use of narcotic drugs
  - Request of not being informed in case of incidental findings Recruitment and screening

Stroke patients are recruited at the stroke unit of the *Hôpital du Valais* (HVS). The member of staff in charge of the recruitment daily checks the list of new entries at the hospital. When a patient is eligible (see Inclusion and Exclusion criteria), the medical staff is consulted, and a first screening visit is organized with the patient. The study is presented in details to the patient, and eligibility is further evaluated. Patients are provided with 24-hours for reflection in regard of participation before signing the consent to participate. If the patient consents, the first visit (T1) is organized during the first week after the stroke, while the patient is most of the time still hospitalized. The procedures are performed in accordance with the ethical approval.

#### Data acquisition and follow-up

The 1^st^ behavioral evaluation and the MRI acquisition are performed at the HVS. The electrophysiological measurements are performed in the laboratory, located in the *Clinique Romande de Réadaptation (CRR)* physically connected to the HVS. The total measurement time is of around 10 hours, distributed in several sessions.

The patients enrolled in the study are then transferred for rehabilitation from the HVS to one of the two rehabilitation clinics collaborating within the present study, that is the CRR and the Berner Klinik (BK; Crans-Montana) or to home. The 3 weeks (T2) behavioral evaluation is performed during the in-patient stay, or in the laboratory if the patient was sent back home after the acute phase. For the 3 months (T3) and 12 months follow-ups (T4), patients are are invited to our laboratory on the HVS/CRR campus for behavioral, MRI and electrophysiological recordings.

### Data management and statistical considerations

Based on previous comparable project (e.g. 41; N=132 patients) and given the estimated feasibility of our extensive multi-modal and multi-domain evaluations, we aimed to recruit up to 100 patients, with a recruitment rate of up to 40 patients a year. As drop-outs are common for this type of longitudinal study, we expect some missing datapoints and will use statistical approaches that are robust to missing values such as linear mixed models.

As the study is mainly explorative in its nature, we do not use a classical power calculation. We will analyze the different behavioral domains individually but we also aim to integrate the multimodal data together in statistical models and computational approaches, in order to determine interactions between the different parameters. Statistics will be performed using either R software (2017, R Core Team, Vienna, https://www.Rproject.org), the SPSS software (2017, IBM SPSS Statistics for Windows, IBM Corp, Armonk, New York), the JASP software, Matlab (v2020b, Mathworks, The MathWorks, Massachusetts, http://www.mathworks.ch) and/or Python (2009, CreateSpace, Scotts Valley, California). We will use a broad spectrum of statistical tools designed for high-dimensional datasets, like general linear model analyses. Besides, we will also use machine learning tools as classifiers, supervised and unsupervised and deep learning algorithms as they provide the opportunity to derive insights from imaging and electrophysiological data coupled with behavior to produce predictive models and to discovering phenotypes of patients (54,55). Specific factors included in statistical modeling will be either hypothesis-driven or mainly exploratory.

## Discussion

As depicted in the introduction, stroke results in multi-domain behavioral deficits in survivors. Although motor deficits (in particular in the upper extremity) are the most impairing, the prevalence of cognitive deficits is also highly important and concerns multiple domains. In addition, they were demonstrated to likely impact the functional recovery and the reintegration in life following stroke, as well as the outcomes of motor rehabilitation (20). Yet, little attention has been paid so far to how cognitive and motor domains are related and influence each other following stroke. Consequently, there is a lack of detailed phenotyping of behavioral outcomes and their evolution though it would be of high interest to improve rehabilitation tailoring (41,56)

Some studies have investigated the relationships between cognitive and motor outcomes (17,22,41,57–59) and showed that cognitive impairments were common even in patients with mild strokes, and that relationships exist between motor and cognitive domains. This highlights the relevance of such multi-domain approaches, emphasizing that motricity and cognition should not be investigated separately. For instance, Einstad and colleagues (2021) have recently demonstrated that poor motor performances are associated with impaired global cognition scores and executive dysfunctions. However, such studies made use of a limited battery of tests and/or focused on one particular timeframe during stroke recovery without any longitudinal assessment (i.e. acute, sub-acute, chronic). Ramsey and colleagues (2017) employed a battery of motor and cognitive tests to evaluate the patients over the course of recovery during the first year; at 1-2 weeks, three months and one year after the stroke. They reported that across multiple domains, sub-acute scores were strong predictors of the performance in the chronic stage and that the magnitude and time course of recovery were comparable between cognitive and motor domains. Specific behavioral clusters were identified (e.g., a strong relationship between motor impairment and attention) and shown as being stable over the three timepoints. In addition, the authors described relationships of interest between domains over the course of recovery (e.g. language deficits influenced the recovery of verbal memory). Interestingly, the authors studied how lesion topography could explain behavior, as it was done in another study from the same group (41) and pointed out that white matter damage could be a key feature in explaining behavioral recovery. Other studies from the same cohort independently investigated the relationships between resting-state fMRI data and behavior by showing that altered functional connectivity correlated with behavioral deficits in the motor and attention domains (60) and in hemi-spatial neglect (61). In addition, the authors demonstrated that memory deficits are better predicted by functional connectivity than by lesion topography while the motor and visual deficits might be better predicted by lesion location than functional connectivity (42). Altogether, these studies emphasized the importance of multi-domain behavioral assessments and the interest of investigating brain-behavior relationships both through structural and functional measures as they provide complementary insights. However, patients enrolled in this cohort were substantially younger than the natural population of stroke survivors (average age 54 ± 11 years old, range 19-83, benchmark 69.2 years in 2005 Greater Cincinnati/Northern Kentucky cohort; 62) and executive functions were not assessed in the battery. Plus, the authors focused on one modality (MRI) to assess brain features which provides rich but limited insights about the neuronal mechanisms underlying post-stroke recovery. To date, no study provided any extensive behavioral evaluation (with an approach centered on the individuals rather than the whole cohort) and during the course of recovery following stroke while combining data with multimodal assessments of brain network plasticity.

A common factor in many of these studies is the interplay between structural and functional connectivity. Structure influences function in the obvious way, while function influences structure in the long term. However, there is strong evidence that the strength of the link between structure and function is domain-dependent. The findings of Siegel, Ramsey and colleagues (2017) suggest that function is tightly coupled to structure in the motor and visual domains, while the two are more decoupled for “higher order” domains such as memory. These findings have been echoed in Preti & Van De Ville’s work (2019), which found that brain regions responsible for “low level sensory function” tend to exhibit strong structural-functional coupling, and vice versa.

The present study aspires to bolster our understanding of mechanics underlying multiple-domain deficits by providing a multi-modal and multi-domain evaluation of stroke patients longitudinally during the first year after the stroke. This research intends to investigate the different behavioral profiles and their dynamics in stroke patients, not only looking at the motor domain but undergoing a holistic approach coupled with neuro-imaging and electrophysiological parameters. Therefore, the originality of the project lies in the multiplicity of the approaches undertaken that will allow a very detailed picture of the recovery and the reorganization in the brain following stroke. Structural, diffusion-weighted and functional MRI will provide the opportunity to study network dysfunctions as well as the complex interactions between brain function and structure. In addition, simultaneous EEG recording during TMS is a promising approach that will enable to explore brain connectivity and recovery pattern for functional networks after stroke by providing a direct measure of the cortical activity induced by TMS. By combining modalities with different advantages (such as either excellent spatial or temporal resolution, structural versus functional information) and by following patients along the first year post-stroke, we will provide a complete dataset allowing to integrate multimodal information in statistical and computational models. The overall goal is to determine interactions between the different parameters as well as factors usable as biomarkers for phenotyping patients in regard of the course and the degree of recovery.

Identifying such biomarkers might help (1) to predict the course of recovery, i.e. to early detect patients that will spontaneously recover and those who will not and, consequently, (2) to personalize the therapeutic strategies in order to meet the individual needs of each patient and to maximize the treatment benefits. Therefore, this work will serve as a basis for improving existing treatments or developing novel and innovative ones tailored to the individual patients’ characteristics by providing a better understanding of neural mechanisms underlying successful recovery. For instance, non-invasive brain stimulation (NIBS) are neuro-technologies that are more and more used in stroke rehabilitation to promote motor recovery (32,63,64) due to their noninvasiveness, relatively low cost and limited side effects. However, there is a high heterogeneity in the outcomes (30,31,55,65,66): effects of NIBS are still limited, which can be partly explained by the use of non-personalized approaches (32,67). Some biomarkers have already been identified to stratify patients in order to assess the individual recovery potential, for instance the cortico-spinal tract integrity as measured by presence or absence of MEP (68). However there is still a lack of fundamental knowledge on the topic especially considering the longitudinal changes in brain dynamics following stroke (23). The detailed phenotyping based on the dataset from the present study might further help to provide extra layers of stratifications allowing more precise predictions about treatment outcomes in order to reduce the number of non-responders (55). Therefore, some potential perspectives are to further design interventional studies to analyze the efficacy of neurotechnologies-based treatment personalized thanks to clustering and stratifying algorithms arising from this research.

Since this work involves plural and extensive multi-modal assessments, it is worthwhile to emphasize that the patients need to be physically and mentally capable of undergoing such multiple recordings. Plus, as the patients need to understand what the project entails, severe language deficits prevent possible participants to be enrolled because they do not have the ability to consent while being transparently informed. Furthermore, the presence of TMS recordings is associated with a consistent list of exclusion criteria related to medication, epilepsy or implants (metallic or electronic) that could interact with the stimulation. These aspects might cause a bias in the recruitment of patients that we need to consider when interpreting the results. Still, we look for cohort as heterogeneous as possible to cluster patients and identify specific patterns of recovery and brain reorganization.

## Summary and conclusions

A better understanding of the neuronal mechanisms associated with recovery-related plasticity and reorganization of the brain networks after a stroke is needed to enhance the understanding of the recovery process, and to predict the outcome and course of recovery. This knowledge will enable to develop and apply interventional strategies in a personalized way to enhance the effects of the treatments for each individual patient. The TiMeS project is a longitudinal, multimodal, and multidomain study of a large, representative cohort of patients during the first year after the stroke, including structural and functional neuro-imaging, electrophysiological and extensive behavioral evaluations. This exploratory research will provide the opportunity to integrate and combine multidimensional data from neuroscience systems methods together with detailed behavioral outcomes to identify specific biomarkers of recovery. This phenotyping will serve as a basis to tailor current rehabilitation strategies according to each patient’s individual needs and to develop innovative personalized neuro-technologies based treatment like NIBS, beyond a one-fits-all approach. Overall, the knowledge gained from this study will pave the way for establishing a close link between basic neuroscience and the development of novel treatments into clinical routine towards precision medicine in stroke, which is highly promising to reduce the burden of the disease.

## Data Availability

All data produced in the present study are available upon reasonable request to the authors

## References

1. GBD 2019 Diseases and Injuries Collaborators. Global burden of 369 diseases and injuries in 204 countries and territories, 1990-2019: a systematic analysis for the Global Burden of Disease Study 2019. Lancet Lond Engl. 17 oct 2020;396(10258):1204⍰22.

2. Gorelick PB. The global burden of stroke: persistent and disabling. Lancet Neurol. 1 mai 2019;18(5):417⍰8.

3. Feigin VL, Krishnamurthi RV, Parmar P, Norrving B, Mensah GA, Bennett DA, et al. Update on the Global Burden of Ischemic and Hemorrhagic Stroke in 1990-2013: The GBD 2013 Study. Neuroepidemiology. 2015;45(3):161⍰76.

4. Hendricks HT, van Limbeek J, Geurts AC, Zwarts MJ. Motor recovery after stroke: A systematic review of the literature. Arch Phys Med Rehabil. 1 nov 2002;83(11):1629⍰37.

5. Kwakkel G, Kollen BJ, van der Grond J, Prevo AJH. Probability of regaining dexterity in the flaccid upper limb: impact of severity of paresis and time since onset in acute stroke. Stroke. sept 2003;34(9):2181⍰6.

6. Lai SM, Studenski S, Duncan PW, Perera S. Persisting consequences of stroke measured by the Stroke Impact Scale. Stroke. juill 2002;33(7):1840⍰4.

7. Langhorne P, Coupar F, Pollock A. Motor recovery after stroke: a systematic review. Lancet Neurol. août 2009;8(8):741⍰54.

8. Barker-Collo S, Feigin VL, Parag V, Lawes CMM, Senior H. Auckland Stroke Outcomes Study: Part 2: Cognition and functional outcomes 5 years poststroke. Neurology. 2 nov 2010;75(18):1608⍰16.

9. Nys GMS, van Zandvoort MJE, de Kort PLM, Jansen BPW, de Haan EHF, Kappelle LJ. Cognitive disorders in acute stroke: prevalence and clinical determinants. Cerebrovasc Dis Basel Switz. 2007;23(5–s6):408⍰16.

10. Barker-Collo S, Feigin V. The impact of neuropsychological deficits on functional stroke outcomes. Neuropsychol Rev. juin 2006;16(2):53⍰64.

11. GBD 2016 Stroke Collaborators. Global, regional, and national burden of stroke, 1990-2016: a systematic analysis for the Global Burden of Disease Study 2016. Lancet Neurol. mai 2019;18(5):439⍰58.

12. Feigin VL, Forouzanfar MH, Krishnamurthi R, Mensah GA. Global burden of stroke: an underestimate Authors’ reply. Lancet Lond Engl. 5 avr 2014;383(9924):1205⍰6.

13. Stinear CM. Prediction of motor recovery after stroke: advances in biomarkers. Lancet Neurol. oct 2017;16(10):826⍰36.

14. Coupar F, Pollock A, Rowe P, Weir C, Langhorne P. Predictors of upper limb recovery after stroke: a systematic review and meta-analysis. Clin Rehabil. 1 avr 2012;26(4):291⍰313.

15. Lang CE, Bland MD, Bailey RR, Schaefer SY, Birkenmeier RL. Assessment of upper extremity impairment, function, and activity after stroke: foundations for clinical decision making. J Hand Ther Off J Am Soc Hand Ther. juin 2013;26(2):104-114;quiz 115.

16. Dennis M, O’Rourke S, Lewis S, Sharpe M, Warlow C. Emotional outcomes after stroke: factors associated with poor outcome. J Neurol Neurosurg Psychiatry. janv 2000;68(1):47⍰52.

17. Ramsey LE, Siegel JS, Lang CE, Strube M, Shulman GL, Corbetta M. Behavioural clusters and predictors of performance during recovery from stroke. Nat Hum Behav. 17 mfévr 2017;1(3):1⍰10.

18. Hochstenbach JB, Anderson PG, van Limbeek J, Mulder TT. Is there a relation between neuropsychologic variables and quality of life after stroke? Arch Phys Med Rehabil. oct 2001;82(10):1360⍰6.

19. Patel MD, Coshall C, Rudd AG, Wolfe CDA. Cognitive impairment after stroke: clinical determinants and its associations with long-term stroke outcomes. J Am Geriatr Soc. avr 2002;50(4):700⍰6.

20. Mullick AA, Subramanian SK, Levin MF. Emerging evidence of the association between cognitive deficits and arm motor recovery after stroke: A meta-analysis. Restor Neurol Neurosci. 1 janv 2015;33(3):389⍰403.

21. Elliott R. Executive functions and their disorders. Br Med Bull. 2003;65:49⍰59.

22. Verstraeten S, Mark RE, Dieleman J, van Rijsbergen M, de Kort P, Sitskoorn MM. Motor Impairment Three Months Post Stroke Implies A Corresponding Cognitive Deficit. J Stroke Cerebrovasc Dis. 1 oct 2020;29(10):105119.

23. Koch P, Schulz R, Hummel FC. Structural connectivity analyses in motor recovery research after stroke. Ann Clin Transl Neurol. mars 2016;3(3):233⍰44.

24. Hartwigsen G. Adaptive Plasticity in the Healthy Language Network: Implications for Language Recovery after Stroke. Neural Plast. 2016;2016:9674790.

25. Barker-Collo S, Feigin V, Lawes C, Senior H, Parag V. Natural history of attention deficits and their influence on functional recovery from acute stages to 6 months after stroke. Neuroepidemiology. 2010;35(4):255⍰62.

26. Byblow WD, Stinear CM, Barber PA, Petoe MA, Ackerley SJ. Proportional recovery after stroke depends on corticomotor integrity. Ann Neurol. déc 2015;78(6):848⍰59.

27. Koch PJ, Hummel FC. Toward precision medicine: tailoring interventional strategies based on noninvasive brain stimulation for motor recovery after stroke. Curr Opin Neurol. août 2017;30(4):388⍰97.

28. Prabhakaran S, Zarahn E, Riley C, Speizer A, Chong JY, Lazar RM, et al. Inter-individual variability in the capacity for motor recovery after ischemic stroke. Neurorehabil Neural Repair. févr 2008;22(1):64⍰71.

29. Marchi NA, Ptak R, Di Pietro M, Schnider A, Guggisberg AG. Principles of proportional recovery after stroke generalize to neglect and aphasia. Eur J Neurol. août 2017;24(8):1084⍰7.

30. Coscia M, Wessel MJ, Chaudary U, Millán JDR, Micera S, Guggisberg A, et al. Neurotechnology-aided interventions for upper limb motor rehabilitation in severe chronic stroke. Brain J Neurol. 1 août 2019;142(8):2182⍰97.

31. Micera S, Caleo M, Chisari C, Hummel FC, Pedrocchi A. Advanced Neurotechnologies for the Restoration of Motor Function. Neuron. 19 2020;105(4):604⍰20.

32. Morishita T, Hummel FC. Non-invasive Brain Stimulation (NIBS) in Motor Recovery After Stroke: Concepts to Increase Efficacy. Curr Behav Neurosci Rep. 1 sept 2017;4(3):280⍰9.

33. Lefaucheur JP, Aleman A, Baeken C, Benninger DH, Brunelin J, Di Lazzaro V, et al. Evidence-based guidelines on the therapeutic use of repetitive transcranial magnetic stimulation (rTMS): An update (2014–2018). Clin Neurophysiol. 1 févr 2020;131(2):474⍰528.

34. Bonkhoff AK, Grefkes C. Precision medicine in stroke: towards personalized outcome predictions using artificial intelligence. Brain. 16 déc 2021;awab439.

35. Koch PJ, Park CH, Girard G, Beanato E, Egger P, Evangelista GG, et al. The structural connectome and motor recovery after stroke: predicting natural recovery. Brain J Neurol. 17 août 2021;144(7):2107⍰19.

36. Guggisberg AG, Koch PJ, Hummel FC, Buetefisch CM. Brain networks and their relevance for stroke rehabilitation. Clin Neurophysiol Off J Int Fed Clin Neurophysiol. 2019;130(7):1098⍰124.

37. Rehme AK, Grefkes C. Cerebral network disorders after stroke: evidence from imaging-based connectivity analyses of active and resting brain states in humans. J Physiol. 1 janv 2013;591(Pt 1):17⍰31.

38. Carrera E, Tononi G. Diaschisis: past, present, future. Brain. 1 sept 2014;137(9):2408⍰22.

39. Grefkes C, Fink GR. Connectivity-based approaches in stroke and recovery of function. Lancet Neurol. 1 févr 2014;13(2):206⍰16.

40. Grefkes C, Ward NS. Cortical Reorganization After Stroke: How Much and How Functional? The Neuroscientist. 1 févr 2014;20(1):56⍰70.

41. Corbetta M, Ramsey L, Callejas A, Baldassarre A, Hacker CD, Siegel JS, et al. Common behavioral clusters and subcortical anatomy in stroke. Neuron. 4 mars 2015;85(5):927⍰41.

42. Siegel JS, Ramsey LE, Snyder AZ, Metcalf NV, Chacko RV, Weinberger K, et al. Disruptions of network connectivity predict impairment in multiple behavioral domains after stroke. Proc Natl Acad Sci U S A. 26 juill 2016;113(30):E4367–4376.

43. Egger P, Evangelista GG, Koch PJ, Park CH, Levin-Gleba L, Girard G, et al. Disconnectomics of the Rich Club Impacts Motor Recovery After Stroke. Stroke. 1 juin 2021;52(6):2115⍰24.

44. Santisteban L, Térémetz M, Bleton JP, Baron JC, Maier MA, Lindberg PG. Upper Limb Outcome Measures Used in Stroke Rehabilitation Studies: A Systematic Literature Review. PLOS ONE. 6 mai 2016;11(5):e0154792.

45. Veldsman M, Brodtmann A. Disconnectomics: Stroke-related disconnection and dysfunction in distributed brain networks. Int J Stroke. 1 janv 2019;14(1):6⍰8.

46. Sporns O. Graph theory methods: applications in brain networks. Dialogues Clin Neurosci. juin 2018;20(2):111⍰21.

47. van den Heuvel MP, Sporns O. Rich-Club Organization of the Human Connectome. J Neurosci. 2 nov 2011;31(44):15775⍰86.

48. Preti MG, Van De Ville D. Decoupling of brain function from structure reveals regional behavioral specialization in humans. Nat Commun. 18 oct 2019;10(1):4747.

49. Kujirai T, Caramia MD, Rothwell JC, Day BL, Thompson PD, Ferbert A, et al. Corticocortical inhibition in human motor cortex. J Physiol. nov 1993;471:501⍰19.

50. Chen R. Interactions between inhibitory and excitatory circuits in the human motor cortex. Exp Brain Res. janv 2004;154(1):1⍰10.

51. Babiloni C, Barry RJ, Başar E, Blinowska KJ, Cichocki A, Drinkenburg WHIM, et al. International Federation of Clinical Neurophysiology (IFCN) EEG research workgroup: Recommendations on frequency and topographic analysis of resting state EEG rhythms. Part 1: Applications in clinical research studies. Clin Neurophysiol. 1 janv 2020;131(1):285⍰307.

52. Tremblay S, Rogasch NC, Premoli I, Blumberger DM, Casarotto S, Chen R, et al. Clinical utility and prospective of TMS-EEG. Clin Neurophysiol Off J Int Fed Clin Neurophysiol. mai 2019;130(5):802⍰44.

53. Sachdev PS, Blacker D, Blazer DG, Ganguli M, Jeste DV, Paulsen JS, et al. Classifying neurocognitive disorders: the DSM-5 approach. Nat Rev Neurol. nov 2014;10(11):634⍰42.

54. Tozlu C, Edwards D, Boes A, Labar D, Tsagaris KZ, Silverstein J, et al. Machine Learning methods predict individual upper limb motor impairment following therapy in chronic stroke. Neurorehabil Neural Repair. mai 2020;34(5):428⍰39.

55. Wessel MJ, Egger P, Hummel FC. Predictive models for response to non-invasive brain stimulation in stroke: A critical review of opportunities and pitfalls. Brain Stimulat. déc 2021;14(6):1456⍰66.

56. Duncan PW, Zorowitz R, Bates B, Choi JY, Glasberg JJ, Graham GD, et al. Management of Adult Stroke Rehabilitation Care: a clinical practice guideline. Stroke. sept 2005;36(9):e100–143.

57. Einstad MS, Saltvedt I, Lydersen S, Ursin MH, Munthe-Kaas R, Ihle-Hansen H, et al. Associations between post-stroke motor and cognitive function: a cross-sectional study. BMC Geriatr. 5 févr 2021;21(1):103.

58. Fong KN, Chan CC, Au DK. Relationship of motor and cognitive abilities to functional performance in stroke rehabilitation. Brain Inj. mai 2001;15(5):443⍰53.

59. Sagnier S, Renou P, Olindo S, Debruxelles S, Poli M, Rouanet F, et al. Gait Change Is Associated with Cognitive Outcome after an Acute Ischemic Stroke. Front Aging Neurosci [Internet]. 2017 [cité 19 janv 2022];9. Disponible sur: https://www.frontiersin.org/article/10.3389/fnagi.2017.00153

60. Siegel JS, Snyder AZ, Ramsey L, Shulman GL, Corbetta M. The effects of hemodynamic lag on functional connectivity and behavior after stroke. J Cereb Blood Flow Metab Off J Int Soc Cereb Blood Flow Metab. déc 2016;36(12):2162⍰76.

61. Ramsey LE, Siegel JS, Baldassarre A, Metcalf NV, Zinn K, Shulman GL, et al. Normalization of network connectivity in hemi-spatial neglect recovery. Ann Neurol. juill 2016;80(1):127⍰41.

62. Kissela BM, Khoury JC, Alwell K, Moomaw CJ, Woo D, Adeoye O, et al. Age at stroke: temporal trends in stroke incidence in a large, biracial population. Neurology. 23 oct 2012;79(17):1781⍰7.

63. Hummel FC, Cohen LG. Non-invasive brain stimulation: a new strategy to improve neurorehabilitation after stroke? Lancet Neurol. août 2006;5(8):708⍰12.

64. Raffin E, Hummel FC. Restoring Motor Functions After Stroke: Multiple Approaches and Opportunities. Neurosci Rev J Bringing Neurobiol Neurol Psychiatry. 2018;24(4):400⍰16.

65. Alia C, Spalletti C, Lai S, Panarese A, Lamola G, Bertolucci F, et al. Neuroplastic Changes Following Brain Ischemia and their Contribution to Stroke Recovery: Novel Approaches in Neurorehabilitation. Front Cell Neurosci. 2017;11:76.

66. Nicolo P, Ptak R, Guggisberg AG. Variability of behavioural responses to transcranial magnetic stimulation: Origins and predictors. Neuropsychologia. juill 2015;74:137⍰44.

67. Grefkes C, Fink GR. Noninvasive brain stimulation after stroke: it is time for large randomized controlled trials! Curr Opin Neurol. déc 2016;29(6):714⍰20.

68. Lindenberg R, Zhu LL, Rüber T, Schlaug G. Predicting functional motor potential in chronic stroke patients using diffusion tensor imaging. Hum Brain Mapp. mai 2012;33(5):1040⍰51.

