## Supplement 1 for "Towards individualized Medicine in Stroke – the TiMeS project: protocol of longitudinal, multi-modal, multi-domain study in stroke"

**Supplement 1 – Neuropsychological evaluation**

Motor and cognitive tests

The battery of tests includes 14 tests assessing the sensory and motor functions, and 26 tests assessing the cognitive functions (see Table 1). It covers the neurocognitive domains described in the DSM-V (Sachdev et al., 2014), i.e. complex attention, language, learning and memory, executive functions, social cognition, perceptual-motor functions, with the latter divided into perceptual and motor functions.

The DSM-V does not name any proprietary tests to objectively assess these functions. Therefore, we selected the tests and questionnaires on the basis of their validity and reproducibility as well as the existence of normative data in the literature. We chose several tests per neurocognitive domain to have an extensive and detailed evaluation, and also to have data that are representative of the several subdomains for each neurocognitive domain.

We use the same battery for each timepoint, excluding a few tests that are skipped during the first timepoint in order to reduce the total time of the evaluation as patients are in general highly fatigable during the first week after the stroke. When possible, versioning was used to reduce learning effects.

The entire evaluation is divided into at least two sessions of 2 to 3 hours per timepoint, depending on the patients’ state and availability. Breaks are imposed to the patients during the session to maintain their attention and concentration, but they are also able to take some rest at any time. The order of the tests is pre-defined to reduce the possible interferences between the different tests, but the examinator keeps to possibility to adjust if this is needed regarding the patients’ state. In any case, the final tests’ order for each evaluation is recorded on the patient’s file.

Specific materials and licenses were acquired for each test. The evaluation is conducted by a neuropsychologist with a relevant clinical experience who follows rigorously the instructions provided by the authors of each test. The instructions of the tests are given, in French, Italian, or German, depending on the patients’ first language. When possible, the version of the tests is adapted to the language used, especially for memory tests for which there are French, Italian, English, and Portuguese versions.

Questionnaires

In addition, 16 questionnaires assess daily life aspects of patients, including physical and mental activity, functional level of dependance and level of reintegration, mental state, fatigue, sleep; etc (see Table 1). The questionnaires are filled by the patients themselves when they are able to, and checked by the neuropsychologist. If needed, the neuropsychologist helps the patients by reading and filling the questionnaires for them. These questionnaires are used at each timepoint plus an additional time between the third and the fourth time points.

| **Test** |  | **Function assessed** |  | **Reference** |
| --- | --- | --- | --- | --- |
| ***Clinical evaluation*** |  |  |  |  |
| NIHSS |  | Neurological examination |  | Brott et al., 1989 |
| ***General cognitive screening*** |  |  |  |  |
| Montreal Cognitive Assessment |  | All cognitive functions – general screening |  | Nasreddine et al., 2005 |
| ***Attention*** |  |  |  |  |
| Test of Attention Performance –  Phasic alert test |  | Alertness, intensity of attention |  | Zimmermann & Fimm, 2016 |
| Test of Attention Performance –  Divided attention test |  | Attentional selectivity, focused attention |  | Zimmermann & Fimm, 2016 |
| D2-R |  | Sustained and focused attention |  | Brickenkamp, 2015 |
| ***Social cognition*** |  |  |  |  |
| Geneva Emotions Recognition Test – Short* |  | Emotion recognition ability |  | Schlegel et al., 2016 |
| ***Executive functions*** |  |  |  |  |
| Frontal Assessment Battery |  | Executive functions |  | Dubois et al., 2000 |
| Stroop Victoria |  | Flexibility, inhibition, information processing speed |  | Spreen & Strauss, 1991; Bayard et al., 2007 |
| Bimanual coordination |  | Planification, programmation |  | Dolivo & Assal, ,1985 |
| Apraxia Screen of Test for Upper-Limb Apraxia |  | Planification, programmation |  | Vanbellingen et al., 2010 |
| CERAD Constructional Praxis |  | Planification, programmation |  | Morris et al., 1989; Roussel & Godefroy, 2016 – GRECOGVASC battery |
| Color Trail Test |  | Planification, programmation, information processing speed | | D'Elia et al., 1996 |
| Bisiach anosognosia scale |  | Self-awareness |  | Bisiach et al., 1986 |
| Somatoparaphrenia test |  | Self-awareness |  | Ronchi et al., adapted |
| 5-points tests* |  | Flexibility |  | Strauss & Knapp, 1982 |
| ***Language*** |  |  |  |  |
| LAST |  | Repetition, denomination, comprehension |  | Flamand-Roze et al., 2011; Koenig-Bruhin et al., 2016 |
| Ardila's language test* |  | Denomination |  | Ardila, 2007 |
| Token Test* |  | Comprehension |  | De Renzi & Vignolo, 1962 |
| Phonological verbal fluency |  | Fluency |  | Roussel & Godefroy, 2016 - GRECOGVASC battery |
| Semantic verbal fluency |  | Fluency |  | Roussel & Godefroy, 2016 - GRECOGVASC battery |
| ***Learning and Memory*** |  |  |  |  |
| Hopkins Verbal Learning Test – revised* |  | Verbal episodic memory |  | Brandt, 1990 |
| Doors test* |  | Visual episodic memory |  | Roussel & Godefroy, 2016 - GRECOGVASC battery |
| Digit span |  | Verbal short-term memory |  | Wechsler, 2008 - Wechsler Adult Intelligence Scale |
| Corsi-Kessels |  | Visual short-term memory |  | Kessels et al., 2000 |
| ***Motor functioning*** |  |  |  |  |
| Fugl-Meyer |  | Upper limb function |  | Fugl-Meyer, 1980 |
| Pinch&Grip |  | Hand strength |  | Mathiowetz et al., 1984 |
| Medical Research Council muscle strength testing |  | Upper limb strength |  | Ciesla et al., 2011, Hislop & Montgomery; 2007 |
| Nine-Hole Peg Test |  | Fine manual dexterity |  | Mathiowetz et al., 1985 |
| Box and Blocks test |  | Gross manual dexterity |  | Mathiowetz et al., 1985 |
| Purdue Pegboard Test |  | Fine manual dexterity, bimanual coordination |  | Tiffin & Asher, 1948 |
| Action Research Arm Test* |  | Upper limb function |  | Lyle, 1981 |
| Modified Ashworth Scale |  | Spasticities |  | Bohannon & Smith, 1987 |
| 2 minutes walk test* |  | Gait |  | Butland et al., 1982 |
| 10 meters walk test* |  | Gait |  | van Hedel et al., 2005 |
| Time Up and Go test* |  | Gait, balance, functional ability |  | Podsiadlo & Richardson, 1991 |
| Berg Balance Scale |  | Balance |  | Berg, 1992 |
| ***Sensory*** |  |  |  |  |
| Rivermead Assessment of Sensory performance |  | Face, hands, feet sensitivity |  | Winward & Halligan, 2002 |
| ***Perceptual function*** |  |  |  |  |
| Overlapping figures test |  | Gnosis, neglect |  | Unilateral neglect assessment battery of the GEREN, 2002 |
| Bisection line test |  | Neglect |  | Unilateral neglect assessment battery of the GEREN, 2002 |
| Bells cancellation test |  | Neglect |  | Unilateral neglect assessment battery of the GEREN, 2002 |
| ***Questionnaires*** |  |  |  |  |
| Stroke Impact Scale (SIS) |  | General recovery |  | Duncan et al., 2003 |
| Hospital Anxiety and Depression Scale (HADS) |  | Anxiety and depression |  | Zigmond & Snaith, 1983 |
| State/Trait Anxiety Inventory for adults (STAI) |  |  |  | Spielberger, 1983 |
| Fear and stress scale |  | Fear and stress |  | Carmen Sandi's version |
| Medical Outcome Study Short Form 12 |  | Reintegration |  | Ware & Sherbourne, 1992; Hurst et al., 1998 |
| Modified Reintegration to Normal Living Index |  |  |  | Wood-Dauphinee et al., 1988 |
| Social Comparison Scale |  | Social comparison |  | Allan & Gilbert, 1995 |
| Generalized Self Efficacy Scale |  | Efficacy |  | Scharzer & Jerusalem, 1995 |
| Pittsburgh Sleep Quality Index |  | Sleep and Fatigue |  | Buysse et al., 1989 |
| Multidimensional Fatigue Inventory |  |  |  | Smets et al., 1995 |
| Feeling of foreigness questionnaire |  | Disembodiement |  |  |
| Neurobehavioral questionnaire |  |  |  |  |
| Barthel index |  | Functional indexes |  | Mahoney & Barthel, 1965 |
| mRS modified Rankin Scale |  |  |  | van Swieten et al., 1988 |
| FAC functional ambulation category |  |  |  | Holden et al., 1984 |
| FIM functional independance |  |  |  | Kidd et al., 1995 |
| Edinburgh handedness inventory |  | Handedness |  | Oldfield, 1971 |

**Table legend** - The table 1. lists all the tests and questionnaires used in TiMeS, with the main function(s) assessed by the tool. The latter information is not exhaustive as some tests could assess functions in multiple neurocognitive domains (for instance, the Color Trail Test mainly assesses some executive functions but could also give information about the sustained attention capacity). The third column indicates the authors / owners of the tests.

**References**

Allan, S., & Gilbert, P. (1995). A social comparison scale : Psychometric properties and relationship to psychopathology. Personality and Individual Differences, 19(3), 293‑299. https://doi.org/10.1016/0191-8869(95)00086-L

Ardila, A. (2007). Toward the development of a cross-linguistic naming test. Archives of Clinical Neuropsychology, 22(3), 297‑307. https://doi.org/10.1016/j.acn.2007.01.016

Azouvi, P., Samuel, C., Louis-Dreyfus, A., Bernati, T., Bartolomeo, P., Beis, J.-M., Chokron, S., Leclercq, M., Marchal, F., Martin, Y., De Montety, G., Olivier, S., Perennou, D., Pradat-Diehl, P., Prairial, C., Rode, G., Siéroff, E., Wiart, L., Rousseaux, M., & French Collaborative Study Group on Assessment of Unilateral Neglect (GEREN/GRECO). (2002). Sensitivity of clinical and behavioural tests of spatial neglect after right hemisphere stroke. Journal of Neurology, Neurosurgery, and Psychiatry, 73(2), 160‑166. https://doi.org/10.1136/jnnp.73.2.160

Bayard, S., Erkes, J., Moroni, C., & Collège des Psychologues Cliniciens spécialisés en Neuropsychologie du Languedoc Roussillon (CPCN Languedoc Roussillon). (2011). Victoria Stroop Test : Normative data in a sample group of older people and the study of their clinical applications in the assessment of inhibition in Alzheimer’s disease. Archives of Clinical Neuropsychology, 26(7), 653‑661. https://doi.org/10.1093/arclin/acr053

Berg, K. O., Maki, B. E., Williams, J. I., Holliday, P. J., & Wood-Dauphinee, S. L. (1992). Clinical and laboratory measures of postural balance in an elderly population. Archives of Physical Medicine and Rehabilitation, 73(11), 1073‑1080.

Bisiach, E., Vallar, G., Perani, D., Papagno, C., & Berti, A. (1986). Unawareness of disease following lesions of the right hemisphere : Anosognosia for hemiplegia and anosognosia for hemianopia. Neuropsychologia, 24(4), 471‑482. https://doi.org/10.1016/0028-3932(86)90092-8

Brickenkamp, R. & Zillmer, E. (1998). The d2 Test of Attention. Seattle, Washington: Hogrefe & Huber Publishers

Bohannon, R. W., & Smith, M. B. (1987). Interrater reliability of a modified Ashworth scale of muscle spasticity. Physical Therapy, 67(2), 206‑207. https://doi.org/10.1093/ptj/67.2.206

Brandt, J. (1991). The hopkins verbal learning test : Development of a new memory test with six equivalent forms. Clinical Neuropsychologist, 5(2), 125‑142. https://doi.org/10.1080/13854049108403297

Brott, T., Adams, H. P., Olinger, C. P., Marler, J. R., Barsan, W. G., Biller, J., Spilker, J., Holleran, R., Eberle, R., & Hertzberg, V. (1989). Measurements of acute cerebral infarction : A clinical examination scale. Stroke, 20(7), 864‑870. https://doi.org/10.1161/01.str.20.7.864

Butland, R. J., Pang, J., Gross, E. R., Woodcock, A. A., & Geddes, D. M. (1982). Two-, six-, and 12-minute walking tests in respiratory disease. British Medical Journal (Clinical research ed.), 284(6329), 1607‑1608.

Buysse, D. J., Reynolds, C. F., Monk, T. H., Berman, S. R., & Kupfer, D. J. (1989). The Pittsburgh sleep quality index : A new instrument for psychiatric practice and research. Psychiatry Research, 28(2), 193‑213. https://doi.org/10.1016/0165-1781(89)90047-4

Ciesla, N., Dinglas, V., Fan, E., Kho, M., Kuramoto, J., & Needham, D. (2011). Manual Muscle Testing : A Method of Measuring Extremity Muscle Strength Applied to Critically Ill Patients. Journal of Visualized Experiments : JoVE, 50, 2632. https://doi.org/10.3791/2632

De Renzi, A., & Vignolo, L. A. (1962). Token test : A sensitive test to detect receptive disturbances in aphasics. Brain: A Journal of Neurology, 85, 665‑678. https://doi.org/10.1093/brain/85.4.665

De Renzi, A., & Vignolo, L. A. (1962). Token test: A sensitive test to detect receptive disturbances in aphasics. Brain: a journal of neurology.

D'Elia, L., & Satz, P. (1989). Color Trails I and 2. Odessa, FL: Psychological Assessment Resources.

Dolivo, C., & Assal, G. (1985). Tests neuropsychologiques rapides pour la recherche d'une détérioration intellectuelle. Psychologie médicale, 17(14), 2093-2095.

Dubois, B., Slachevsky, A., Litvan, I., & Pillon, B. (2000). The FAB : A frontal assessment battery at bedside. Neurology, 55(11), 1621‑1626. https://doi.org/10.1212/WNL.55.11.1621

Duncan, P. W., Lai, S. M., Bode, R. K., Perera, S., & DeRosa, J. (2003). Stroke Impact Scale-16 : A brief assessment of physical function. Neurology, 60(2), 291‑296. https://doi.org/10.1212/01.wnl.0000041493.65665.d6

Flamand-Roze, C., Falissard, B., Roze, E., Maintigneux, L., Beziz, J., Chacon, A., Join-Lambert, C., Adams, D., & Denier, C. (2011). Validation of a new language screening tool for patients with acute stroke : The Language Screening Test (LAST). Stroke, 42(5), 1224‑1229. https://doi.org/10.1161/STROKEAHA.110.609503

Förderreuther, S., Sailer, U., & Straube, A. (2004). Impaired self-perception of the hand in complex regional pain syndrome (CRPS). Pain, 110(3), 756‑761. https://doi.org/10.1016/j.pain.2004.05.019

Fugl-Meyer, A. R. (1980). Post-stroke hemiplegia assessment of physical properties. Scandinavian Journal of Rehabilitation Medicine. Supplement, 7, 85‑93.

Galer, B. S., & Jensen, M. (1999). Neglect-like symptoms in complex regional pain syndrome : Results of a self-administered survey. Journal of Pain and Symptom Management, 18(3), 213‑217. https://doi.org/10.1016/s0885-3924(99)00076-7

Godefroy, O., Leclercq, C., Roussel, M., Moroni, C., Quaglino, V., Beaunieux, H., Tallia, H., Nédélec-Ciceri, C., Bonnin, C., Thomas-Anterion, C., Varvat, J., Aboulafia-Brakha, T., Assal, F., & GRECOG-VASC Neuropsychological Committee. (2012). French adaptation of the vascular cognitive impairment harmonization standards : The GRECOG-VASC study. International Journal of Stroke: Official Journal of the International Stroke Society, 7(4), 362‑363. https://doi.org/10.1111/j.1747-4949.2012.00794.x

Holden, M. K., Gill, K. M., Magliozzi, M. R., Nathan, J., & Piehl-Baker, L. (1984). Clinical gait assessment in the neurologically impaired. Reliability and meaningfulness. Physical Therapy, 64(1), 35‑40. https://doi.org/10.1093/ptj/64.1.35

Hurst, N. P., Ruta, D. A., & Kind, P. (1998). Comparison of the MOS short form-12 (SF12) health status questionnaire with the SF36 in patients with rheumatoid arthritis. British Journal of Rheumatology, 37(8), 862‑869. https://doi.org/10.1093/rheumatology/37.8.862

Hislop, H. J., & Montgomery, J. (2007). Daniels and Worthingham’s Muscle Testing Techniques of Manual Examination, Eight Edition. Missouri: Saunders Elsevier, 95-98.

Jerusalem, M., & Schwarzer, R. (1995). General Self-Efficacy Scale--Revised--English Version. APA PsycTests.

Kessels, R. P. C., van Zandvoort, M. J. E., Postma, A., Kappelle, L. J., & de Haan, E. H. F. (2000). The Corsi Block-Tapping Task : Standardization and Normative Data. Applied Neuropsychology, 7(4), 252‑258. https://doi.org/10.1207/S15324826AN0704_8

Kidd, D., Stewart, G., Baldry, J., Johnson, J., Rossiter, D., Petruckevitch, A., & Thompson, A. J. (1995). The Functional Independence Measure : A comparative validity and reliability study. Disability and Rehabilitation, 17(1), 10‑14. https://doi.org/10.3109/09638289509166622

Koenig-Bruhin, M., Vanbellingen, T., Schumacher, R., Pflugshaupt, T., Annoni, J. M., Müri, R. M., Bohlhalter, S., & Nyffeler, T. (2016). Screening for Language Disorders in Stroke : German Validation of the Language Screening Test (LAST). Cerebrovascular Diseases Extra, 6(1), 27‑31. https://doi.org/10.1159/000445778

Luszczynska, A., Scholz, U., & Schwarzer, R. (2005). The general self-efficacy scale : Multicultural validation studies. The Journal of Psychology, 139(5), 439‑457. https://doi.org/10.3200/JRLP.139.5.439-457

Lyle, R. C. (1981). A performance test for assessment of upper limb function in physical rehabilitation treatment and research. International Journal of Rehabilitation Research, 4(4), 483‑492.

Mahoney, F. I., & Barthel, D. W. (1965). FUNCTIONAL EVALUATION : THE BARTHEL INDEX. Maryland State Medical Journal, 14, 61‑65.

Mathiowetz, V., Volland, G., Kashman, N., & Weber, K. (1985). Adult norms for the Box and Block Test of manual dexterity. The American Journal of Occupational Therapy: Official Publication of the American Occupational Therapy Association, 39(6), 386‑391. https://doi.org/10.5014/ajot.39.6.386

Mathiowetz, V., Weber, K., Kashman, N., & Volland, G. (1985). Adult Norms for the Nine Hole Peg Test of Finger Dexterity. The Occupational Therapy Journal of Research, 5(1), 24‑38. https://doi.org/10.1177/153944928500500102

Mathiowetz, V., Weber, K., Volland, G., & Kashman, N. (1984). Reliability and validity of grip and pinch strength evaluations. The Journal of Hand Surgery, 9(2), 222‑226. https://doi.org/10.1016/s0363-5023(84)80146-x

Morris, J. C., Heyman, A., Mohs, R. C., Hughes, J. P., van Belle, G., Fillenbaum, G. D. M. E., ... & Clark, C. (1989). The consortium to establish a registry for Alzheimer's disease (CERAD): I. Clinical and neuropsychological assessment of Alzheimer's disease. Neurology.

Nasreddine, Z. S., Phillips, N. A., Bédirian, V., Charbonneau, S., Whitehead, V., Collin, I., Cummings, J. L., & Chertkow, H. (2005). The Montreal Cognitive Assessment, MoCA : A Brief Screening Tool For Mild Cognitive Impairment. Journal of the American Geriatrics Society, 53(4), 695‑699. https://doi.org/10.1111/j.1532-5415.2005.53221.x

Oldfield, R. C. (1971). The assessment and analysis of handedness : The Edinburgh inventory. Neuropsychologia, 9(1), 97‑113. https://doi.org/10.1016/0028-3932(71)90067-4

Podsiadlo, D., & Richardson, S. (1991). The timed « Up & Go » : A test of basic functional mobility for frail elderly persons. Journal of the American Geriatrics Society, 39(2), 142‑148. https://doi.org/10.1111/j.1532-5415.1991.tb01616.x

Regard, M., Strauss, E., & Knapp, P. (1982). Children’s Production on Verbal and Non-Verbal Fluency Tasks. Perceptual and Motor Skills, 55(3), 839‑844. https://doi.org/10.2466/pms.1982.55.3.839

Roussel, M., & Godefroy, O. (2016). La batterie GRECOGVASC: Evaluation et diagnostic des troubles neurocognitifs vasculaires avec ou sans contexte d'accident vasculaire cérébral. De Boeck Supérieur.

Sandi, C. (2013). Stress and cognition. WIREs Cognitive Science, 4(3), 245‑261. https://doi.org/10.1002/wcs.1222

Schlegel, K., Grandjean, D., & Scherer, K. R. (2014). Introducing the Geneva emotion recognition test : An example of Rasch-based test development. Psychological Assessment, 26(2), 666‑672. https://doi.org/10.1037/a0035246

Smets, E. M. A., Garssen, B., Bonke, B., & De Haes, J. C. J. M. (1995). The multidimensional Fatigue Inventory (MFI) psychometric qualities of an instrument to assess fatigue. Journal of Psychosomatic Research, 39(3), 315‑325. https://doi.org/10.1016/0022-3999(94)00125-O

Snaith, R. P. (2003). The Hospital Anxiety And Depression Scale. Health and Quality of Life Outcomes, 1(1), 29. https://doi.org/10.1186/1477-7525-1-29

Spielberger, C. D. (1983). State-trait anxiety inventory for adults (STAI-AD). APA PsycTests.

Spielberger, C. D. (2010). State-Trait Anxiety Inventory. In The Corsini Encyclopedia of Psychology (p. 1‑1). John Wiley & Sons, Ltd. https://doi.org/10.1002/9780470479216.corpsy0943

Strauss, E., Strauss, P. of P. E., Sherman, N. and A. A. P. D. of P. and C. N. E. M. S., Sherman, E. M. S., Spreen, O., & Spreen, B. P. of P. O. (2006). A Compendium of Neuropsychological Tests : Administration, Norms, and Commentary. Oxford University Press.

Tiffin, J., & Asher, E. J. (1948). The Purdue Pegboard : Norms and studies of reliability and validity. Journal of Applied Psychology, 32(3), 234‑247. https://doi.org/10.1037/h0061266

Vallar, G., & Ronchi, R. (2009). Somatoparaphrenia : A body delusion. A review of the neuropsychological literature. Experimental Brain Research, 192(3), 533‑551. https://doi.org/10.1007/s00221-008-1562-y

Vanbellingen, T., Kersten, B., Winckel, A. V. de, Bellion, M., Baronti, F., Müri, R., & Bohlhalter, S. (2011). A new bedside test of gestures in stroke : The apraxia screen of TULIA (AST). Journal of Neurology, Neurosurgery & Psychiatry, 82(4), 389‑392. https://doi.org/10.1136/jnnp.2010.213371

van Hedel, H. J., Wirz, M., & Dietz, V. (2005). Assessing walking ability in subjects with spinal cord injury : Validity and reliability of 3 walking tests. Archives of Physical Medicine and Rehabilitation, 86(2), 190‑196. https://doi.org/10.1016/j.apmr.2004.02.010

van Swieten, J. C., Koudstaal, P. J., Visser, M. C., Schouten, H. J., & van Gijn, J. (1988). Interobserver agreement for the assessment of handicap in stroke patients. Stroke, 19(5), 604‑607. https://doi.org/10.1161/01.str.19.5.604

Wechsler, D. (1955). Wechsler adult intelligence scale--. Archives of Clinical Neuropsychology.

Ware, J. E., & Sherbourne, C. D. (1992). The MOS 36-Item Short-Form Health Survey (SF-36) : I. Conceptual Framework and Item Selection. Medical Care, 30(6), 473‑483.

Winward, C. E., Halligan, P. W., & Wade, D. T. (2002). The Rivermead Assessment of Somatosensory Performance (RASP) : Standardization and reliability data. Clinical Rehabilitation, 16(5), 523‑533. https://doi.org/10.1191/0269215502cr522oa

Wood-Dauphinee, S. L., Opzoomer, M. A., Williams, J. I., Marchand, B., & Spitzer, W. O. (1988). Assessment of global function : The reintegration to normal living index. Archives of Physical Medicine and Rehabilitation, 69(8), 583‑590. Scopus.

Zimmermann, P., & Fimm, B. (2002). A test battery for attentional performance. Applied neuropsychology of attention. Theory, diagnosis and rehabilitation, 110-151.
