## Supplement 2 for "Towards individualized Medicine in Stroke – the TiMeS project: protocol of longitudinal, multi-modal, multi-domain study in stroke"

**Supplement 2 – Neuroimaging recordings**

***Data acquisition***

Structural, functional, diffusion-weighted and susceptibility-weighted imaging data are acquired using a 3T MAGNETOM Prisma scanner (Siemens, Erlangen, Germany) with a 64-channel head and neck coil.

Diffusion-weighted imaging (DWI)

Diffusion-weighted images are acquired using pulsed gradient spin echo technique (TR = 5000 ms; TE = 77 ms; slices = 84; FOV = 234x234 mm; voxel resolution = 1.6 × 1.6 × 1.6 mm^3^; readout bandwidth = 1630 Hz/pixels; GRAPPA acceleration factor = 3). Seven T2-weighted images without diffusion weighting (b = 0 s/mm^2^) are acquired, including one in opposite phase encoded direction. Further, 101 images with noncollinear diffusion gradient directions distributed uniformly over the half-sphere covering 5 diffusion gradient strengths are measured (b-values = [300, 700, 1000, 2000, 3000] s/mm^2^; shell-samples = [3, 7, 16, 29, 46]). Total acquisition time is 11:06 min.

T1-weighted image

A T1-weighted image is acquired using a 3D Magnetization-Prepared Rapid Gradient-Echo sequence (MPRAGE, TR = 2300 ms; TE = 2.96 ms; flip angle = 9°; slices = 192; voxel size =1 × 1 × 1 mm, FOV 256 × 256 mm, acquisition time = 5:12min).

multi-echo GRASE

A multi-echo T_2_ GRASE sequence supporting CAIPIRINHA parallel imaging (Piredda et al., 2021) is used for myelin water fraction and multi-compartment T2 imaging (in-plane resolution 1.6mm x 1.6mm; slice thickness 1.6mm; 84 slices; acquisition time = 10:30min)..

Blood oxygenation-level dependent (BOLD) MRI / functional MRI (fMRI)

Resting-state BOLD data (with fixation cross) is acquired using a multi-band echo-planar imaging (EPI) sequence. In total, 385 functional volumes are acquired and every volume comprises 75 axial slices covering the whole brain (in-plane resolution = 2 mm x 2 mm; slice thickness 2 mm; no gap, FOV = 256mm, TE = 32 ms, TR=1250 ms, flip angle = 58°, Accel. Factor slice = 5, acquisition time = 8:13min).

GRE field mapping

GRE field mapping is acquired covering the whole brain (75 axial slices), using the following imaging parameters: in-plane resolution = 2mm x 2mm; slice thickness = 2mm; FOV = 224 x 224 mm², TR = 704 mm TE1 = 4.92 ms, TE2 = 7.38 ms , flip angle = 60°, acquisition time = 2:41min.

mp2rage

3D T1-weighted Magnetization-Prepared 2 Rapid Gradient-Echo (MP2RAGE) sequence (INV1 or TI1 = 700 ms, flip angle = 4° and INV2 or TI2 = 2500 ms, flip angle = 5°, TE = 2.98 ms, TR = 5000 ms, acquisition time = 8:22min) with an isotropic voxel resolution of 1 mm^3^.

Susceptibility-weighted imaging (SWI)

We are using a susceptibility-weighed imaging sequence with a TR of 28 ms, a TE of 20ms, FOV = 230 x 230 mm², FOV phase = 78.1%, in-plane resolution of 0.6mm x 0.6mm, slice thickness = 1.2mm, flip angle = 15°, acquisition time = 4:07min.

***Image Analysis***

Lesion segmentation

All the lesion masks were hand-drawn using mrview from MRtrix3 (Tournier et al., 2019) and subsequently verified by a neurologist.

Multi-echo T_2_ imaging

The multi-echo T_2_ data is filtered using a 3D total variation algorithm before fitting (*denoise-tv-chambolle* function of the scikit-image python toolbox (Walt et al., 2014). The data is then registered to the T1-weighted image using using FSL FLIRT (Jenkinson and Smith, 2001) and FNIRT (Andersson et al., 2007; Jenkinson et al., 2012) methods. The myelin water fraction and multi-compartment T_2_ maps are obtained using the L-curve-I method (Canales-Rodríguez et al., 2021) available at https://github.com/ejcanalesr/multicomponent-T2-toolbox.

Diffusion-weighted imaging

The diffusion-weighted images are preprocessed using MRtrix3 (Tournier et al., 2019), FSL (Smith et al., 2004), and Dipy (Garyfallidis et al., 2014). First, Gibbs ringing artefacts are removed from the data (Kellner et al., 2016), then motion artefact reduction, as well as field inhomogeneity, susceptibility-induced off-resonance field and eddy currents correction are performed using FSL TOPUP and EDDY (Andersson et al., 2003; Andersson & Sotiropoulos, 2016). Diffusion-weighted images were then corrected for spatial intensity variations (Zhang et al., 2001). Multi-shell multi-tissue constrained spherical deconvolution (Jeurissen et al., 2014) is used to estimate the fibre orientation distributions within each voxel. Whole-brain probabilistic tractography is performed using the MRtrix3 second-order integration over fibre orientation distribution (iFOD2) algorithm, initiating streamlines in all voxels of the white matter. For each dataset, 1 million streamlines are selected with both endpoints in the individual cortical or subcortical mask using the Dipy software package (Garyfallidis et al., 2014). The obtained tractograms are weighted fitting the underlying diffusion compartment model using a Stick-Ball-Zeppelin model based on COMMIT (Daducci et al., 2015). The stick compartment models the intra-axonal water with parallel diffusivity of 1.7 µm^2^/ms and no perpendicular diffusivity. The Ball compartment models the extra-axonal water with isotropic diffusivity of 1.7 µm^2^/ms and free water with diffusivity of 3.0 µm^2^/ms (Alexander, 2008; Scholz et al., 2009). The Zeppelin compartment models of the extra-axonal water with parallel diffusivity of 1.7 µm^2^/ms and perpendicular diffusivity of 0.51 µm^2^/ms (Alexander, 2008). Tissue partial volume estimates are obtained from the T1-weighted image using the FSL FAST (Zhang et al., 2001) and BET (Smith, 2002) methods. The T1-weighted image is registered to the average b0 image using FSL FLIRT (Jenkinson & Smith, 2001) and FNIRT (Andersson et al., 2007; Jenkinson et al., 2012) methods.

For the cortical parcellation, we choose either the Destrieux (74 areas per hemisphere) (Destrieux et al., 2010) or the Glasser atlas (180 areas per hemisphere) (Glasser et al., 2016) using FreeSurfer (Fischl, 2012; Fischl et al., 2004). To the Destrieux parcellation, we add subcortical areas (thalamus, caudate, putamen. hippocampus, amygdala), the cerebellum ) and a subdivision of the brainstem (midbrain, pons, medulla), totalizing 163 cortical and subcortical areas using the Destrieux cortical parcellation. The parcellations are performed on the T1-weighted image. For stroke patients, the voxels corresponding to the lesion are stamped out and replaced by the mirrored voxels of the contralateral side. For each participant, a structural connectome (SC) is built with 163 (Destrieux atlas), respectively 360 (Glasser atlas), pairs of areas obtained through the parcellation.

Functional (BOLD) Imaging

All preprocessing and statistical analyses are conducted using the SPM12 package (Wellcome Department of Cognitive Neurology, London, UK; www.fil.ion.ucl.ac.uk/spm) running on MATLAB (v2020b, Mathworks, The MathWorks, Massachusetts, http://www.mathworks.ch). Functional images are realigned to the mean functional image. Then, the anatomical image is co-registered to the mean functional image. The anatomical image is segmented into tissue maps based on tissue probability maps of SPM12 (for patients, voxels corresponding to the lesion are not considered). The resulting forward deformation field is used to warp both the anatomical and functional images into MNI space. Finally, the functional images are smoothed using a Gaussian kernel (FWHM = 6 mm). The first 10 volumes are discarded so that the fMRI signal achieves steady-state magnetization, resulting in 375 functional volumes.

Using the conn toolbox (Whitfield-Gabrieli & Nieto-Castanon, 2012), voxel fMRI time courses are detrended and nuisance variables are regressed out (6 head motion parameters, average cerebrospinal fluid and white matter signal). Finally, a band-pass filter is applied (0.01-0.15Hz) to improve signal-to-noise ratio. The average timeseries is then extracted for every ROI of the Glasser parcellation and a functional connectome is obtained by computing the Pearson correlation between timeseries.

References

Alexander, D. C. (2008). A general framework for experiment design in diffusion MRI and its application in measuring direct tissue-microstructure features. *Magnetic Resonance in Medicine*, *60*(2), 439‑448. https://doi.org/10.1002/mrm.21646

Andersson, J. L. R., Skare, S., & Ashburner, J. (2003). How to correct susceptibility distortions in spin-echo echo-planar images : Application to diffusion tensor imaging. *NeuroImage*, *20*(2), 870‑888. https://doi.org/10.1016/S1053-8119(03)00336-7

Andersson, J. L., Jenkinson, M., & Smith, S. (2007). Non-linear registration, aka Spatial normalisation FMRIB technical report TR07JA2. *FMRIB Analysis Group of the University of Oxford*, *2*(1), e21.

Andersson, J. L. R., & Sotiropoulos, S. N. (2016). An integrated approach to correction for off-resonance effects and subject movement in diffusion MR imaging. *NeuroImage*, *125*, 1063‑1078. https://doi.org/10.1016/j.neuroimage.2015.10.019

Ashburner, J., Barnes, G., Chen, C. C., Daunizeau, J., Flandin, G., Friston, K., ... & Penny, W. (2014). SPM12 manual. *Wellcome Trust Centre for Neuroimaging, London, UK*, *2464*.

Jeurissen, B., Tournier, JB., Dhollander, T., Connelly, A., & Sijbers, J.. (2014). Multi-tissue constrained spherical deconvolution for improved analysis of multi-shell diffusion MRI data. *NeuroImage*, *103*, 411‑426. https://doi.org/10.1016/J.NEUROIMAGE.2014.07.061

Canales-Rodríguez, E. J., Pizzolato, M., Piredda, G. F., Hilbert, T., Kunz, N., Pot, C., Yu, T., Salvador, R., Pomarol-Clotet, E., Kober, T., Thiran, J.-P., & Daducci, A. (2021). Comparison of non-parametric T2 relaxometry methods for myelin water quantification. *Medical Image Analysis*, *69*, 101959. https://doi.org/10.1016/j.media.2021.101959

Daducci, A., Dal Palu, A., Lemkaddem, A., & Thiran, J.-P. (2015). COMMIT: Convex Optimization Modeling for Microstructure Informed Tractography. *IEEE Transactions on Medical Imaging*, *34*(1), 246‑257. https://doi.org/10.1109/TMI.2014.2352414

Destrieux, C., Fischl, B., Dale, A., & Halgren, E. (2010). Automatic parcellation of human cortical gyri and sulci using standard anatomical nomenclature. *NeuroImage*, *53*(1), 1‑15. https://doi.org/10.1016/j.neuroimage.2010.06.010

Kellner, E., Dhital, B., Kiselev, VG., & Reisert, M. (2016). Gibbs-ringing artifact removal based on local subvoxel-shifts. *Magnetic resonance in medicine*, *76*(5), 1574‑1581. https://doi.org/10.1002/MRM.26054

Fischl, B. (2012). FreeSurfer. *NeuroImage*, *62*(2), 774‑781. https://doi.org/10.1016/j.neuroimage.2012.01.021

Fischl, B., van der Kouwe, A., Destrieux, C., Halgren, E., Ségonne, F., Salat, D. H., Busa, E., Seidman, L. J., Goldstein, J., Kennedy, D., Caviness, V., Makris, N., Rosen, B., & Dale, A. M. (2004). Automatically parcellating the human cerebral cortex. *Cerebral Cortex (New York, N.Y.: 1991)*, *14*(1), 11‑22. https://doi.org/10.1093/cercor/bhg087

Garyfallidis, E., Brett, M., Amirbekian, B., Rokem, A., van der Walt, S., Descoteaux, M., Nimmo-Smith, I., & Dipy Contributors. (2014). Dipy, a library for the analysis of diffusion MRI data. *Frontiers in Neuroinformatics*, *8*, 8. https://doi.org/10.3389/fninf.2014.00008

Glasser, M. F., Coalson, T. S., Robinson, E. C., Hacker, C. D., Harwell, J., Yacoub, E., ... & Van Essen, D. C. (2016). A multi-modal parcellation of human cerebral cortex. *Nature*, *536*(7615), 171-178.

Jenkinson, M., Beckmann, C. F., Behrens, T. E. J., Woolrich, M. W., & Smith, S. M. (2012). FSL. *NeuroImage*, *62*(2), 782‑790. https://doi.org/10.1016/j.neuroimage.2011.09.015

Jenkinson, M., & Smith, S. (2001). A global optimisation method for robust affine registration of brain images. *Medical Image Analysis*, *5*(2), 143‑156. https://doi.org/10.1016/s1361-8415(01)00036-6

Jeurissen, B., Tournier, J.-D., Dhollander, T., Connelly, A., & Sijbers, J. (2014). Multi-tissue constrained spherical deconvolution for improved analysis of multi-shell diffusion MRI data. *NeuroImage*, *103*, 411‑426. https://doi.org/10.1016/j.neuroimage.2014.07.061

Kellner, E., Dhital, B., Kiselev, V. G., & Reisert, M. (2016). Gibbs-ringing artifact removal based on local subvoxel-shifts. *Magnetic Resonance in Medicine*, *76*(5), 1574‑1581. https://doi.org/10.1002/mrm.26054

Piredda, G. F., Hilbert, T., Thiran, J.-P., & Kober, T. (2021). Probing myelin content of the human brain with MRI : A review. *Magnetic Resonance in Medicine*, *85*(2), 627‑652. https://doi.org/10.1002/mrm.28509

Scholz, J., Klein, M. C., Behrens, T. E. J., & Johansen-Berg, H. (2009). Training induces changes in white-matter architecture. *Nature Neuroscience*, *12*(11), 1370‑1371. https://doi.org/10.1038/nn.2412

Smith, S. M. (2002). Fast robust automated brain extraction. *Human Brain Mapping*, *17*(3), 143‑155. https://doi.org/10.1002/hbm.10062

Tournier, J.-D., Smith, R., Raffelt, D., Tabbara, R., Dhollander, T., Pietsch, M., Christiaens, D., Jeurissen, B., Yeh, C.-H., & Connelly, A. (2019). MRtrix3 : A fast, flexible and open software framework for medical image processing and visualisation. *NeuroImage*, *202*, 116137. https://doi.org/10.1016/j.neuroimage.2019.116137

Walt, S. van der, Schönberger, J. L., Nunez-Iglesias, J., Boulogne, F., Warner, J. D., Yager, N., Gouillart, E., & Yu, T. (2014). scikit-image : Image processing in Python. *PeerJ*, *2*, e453. https://doi.org/10.7717/peerj.453

Whitfield-Gabrieli, S., & Nieto-Castanon, A. (2012). Conn : A functional connectivity toolbox for correlated and anticorrelated brain networks. *Brain Connectivity*, *2*(3), 125‑141. https://doi.org/10.1089/brain.2012.0073

Zhang, Y., Brady, M., & Smith, S. (2001). Segmentation of brain MR images through a hidden Markov random field model and the expectation-maximization algorithm. *IEEE Transactions on Medical Imaging*, *20*(1), 45‑57. https://doi.org/10.1109/42.906424
