## Supplementary material for "Towards individualized Medicine in Stroke – the TiMeS project: protocol of longitudinal, multi-modal, multi-domain study in stroke": Suplement 3

**Supplement 3 – Electrophysiological recordings**

Electrophysiological recordings (resting-state EEG and TMS-EEG coupling) are performed at each time point post stroke: within a week, three weeks, three months, and one year after the ictal event; not for all patients the 4 timepoints are available.

Electroencephalography (EEG)

EEG recordings are acquired using a 64 passive electrodes EEG BrainCap-MR compatible with TMS (Brain Vision LLC, North Carolina, USA) with the reference electrode at FCz and the ground at AFz. The data is recorded with the help of BrainVision Recorder (Brain Vision LLC, North Carolina, USA). The experiment is performed in a faraday cage (IAC Acoustics, Illinois, USA) to limit power line interference. Each electrode is brought to an impedance below 5 kOhm or as low as possible. Impedance levels are recorded at the beginning and end of the session. Data are recorded using DC mode, a resolution of 0.5 μV and a low-pass filter (cutoff frequency of 1 kHz) at a sampling rate of 5 kHz.

Electromyography (EMG)

The EMG activity is recorded using pairs of disposable Ag-AgCl electrodes on 7 muscles in the affected side (First Dorsal Interossei, FDI; Abductor Digiti Minimi, ADM; Abductor Pollicis Brevis, APB; Flexor Carpi Ulnaris, FCU; Flexor Carpi Radialis, FCR; Extensor Carpi Radialis, ECR; Extensor Carpi Ulnaris, ECU) and 1 muscle on the non-affected one (FDI). The signal is amplified and sampled at 3 kHz using a Noraxon DTS Receiver (Scottasdale, Arizona, United States) using a band-pass filter from 10 Hz to 1000 Hz, and finally fed to the Signal software (Cambridge Electronic Design Limited, Cambridge, UK) for further processing.

Transcranial Magnetic Stimulation

Neuronavigated TMS is applied using a MagPro X100 stimulator connected to an MC-B70 coil (Magventure, Farum, Denmark). A neuronavigation system (Localite GmbH, Bonn, Germany) is used throughout the experiment to track and record the position of the stimulation coil in respect to the patient’s individual anatomy, using T1-weighted images (see supp. 2). EEG channel coordinates are also recorded using the neuronavigation system. Biphasic pulses inducing a posterior to anterior current direction are delivered over the first dorsal intraosseous (FDI) hotspot of the affected arm. The stimulation intensity is adjusted to produce MEPs presenting a peak-to-peak amplitude between 0.5 to 1 mV. If no visible MEP (50 µV) can be elicited at maximal stimulator output, the intensity is set similarly on the unaffected hemisphere. The resting motor threshold (rMT) is defined as the lowest intensity necessary to evoke MEPs higher than 50 µV in at least 5 out of 10 trials. Two types of stimulation are applied: a single pulse at the supra-motor threshold intensity fixed earlier or a double pulse (Short-interval Cortical Inhibition; SICI) comprised of a conditioning pulse at 80% rMT followed by a test pulse at the supra-motor threshold intensity, with an inter-stimulus interval of 3 ms.

In order to reduce electromagnetic and acoustic interference resulting from the TMS, electrodes wires are oriented perpendicular to the magnetic field, athin layer of foam is applied between the coil and the scalp and white noise is played through earplugs at a volume covering the sound of the TMS or as loud as tolerated (ter Braack et al., 2015; Veniero et al., 2009).

Data processing

EMG: EMG data are exported to Matlab files to be used with a custom graphical interface for pre-processing. Rejection criteria are as follows: trials with muscle pre-activation exceeding ± 25 μV from baseline less than 100 ms before TMS onset (Delorme & Makeig, 2004) and/or ± 100 μV from baseline 500–100 ms before the pulse are rejected. Trials containing artefacts or with documented suboptimal coil placement are rejected from further analysis. The main features of interest consist of MEP peak-to-peak amplitudes and latencies.

EEG: EEG data are analyzed on Matlab (MathWorks, Massachusetts, USA) using the Fieldtrip (Oostenveld et al., 2011), Brainstorm (Tadel et al., 2011), EEGLAB (Delorme & Makeig, 2004) and TESA (Rogasch et al., 2017) toolboxes. Resting-state EEG recording are preprocessed following international standards (Babiloni et al., 2020) that were successfully used in previous studies on stroke patients (Snyder et al., 2021). First, the continuous data are epoched in non-overlapping 2 s time windows. After removing bad channels and trials, an ICA is performed in order to filter out any remaining ocular, muscular or electrical artifacts. Data are finally re-referenced (average reference), and time-frequency maps are drawn from them by means of multitaper frequency transformation within the 1-50 Hz frequency bandwidth. The asymmetry indices (Snyder et al., 2021) and the brain oscillatory modes drawn from tensor decomposition methods (Tangwiriyasakul et al., 2019) are the main outcomes of interest for this analysis.

Regarding TMS-EEG recordings, the preprocessing pipeline is similar to the one defined by Rogasch et al. (2017) and consists of: epoching, removing data corrupted by the TMS pulse [-5,+20ms], removing bad trials and channels after a visual inspection, removing the remaining TMS artefact and others artefacts such as eye blinks or large muscle artefacts using two rounds of ICA,and finally re-referencing to the average reference. TMS evoked potentials (TEPs) and induced oscillations are then computed by averaging the signal in the time and time-frequency domains respectively. These features allow for the study of both local properties of the stimulated tissue, such as cortical excitability (Raffin et al., 2020), and large-scale properties of the stimulated brain, such as functional and effective connectivity (Tremblay et al., 2019).

**References**

Babiloni, C., Barry, R. J., Başar, E., Blinowska, K. J., Cichocki, A., Drinkenburg, W. H. I. M., Klimesch, W., Knight, R. T., Lopes da Silva, F., Nunez, P., Oostenveld, R., Jeong, J., Pascual-Marqui, R., Valdes-Sosa, P., & Hallett, M. (2020). International Federation of Clinical Neurophysiology (IFCN) – EEG research workgroup : Recommendations on frequency and topographic analysis of resting state EEG rhythms. Part 1: Applications in clinical research studies. *Clinical Neurophysiology*, *131*(1), 285‑307. https://doi.org/10.1016/j.clinph.2019.06.234

Delorme, A., & Makeig, S. (2004). EEGLAB : An open source toolbox for analysis of single-trial EEG dynamics including independent component analysis. *Journal of Neuroscience Methods*, *134*(1), 9‑21. https://doi.org/10.1016/j.jneumeth.2003.10.009

Oostenveld, R., Fries, P., Maris, E., & Schoffelen, J.-M. (2011). FieldTrip : Open source software for advanced analysis of MEG, EEG, and invasive electrophysiological data. *Computational Intelligence and Neuroscience*, *2011*, 156869. https://doi.org/10.1155/2011/156869

Raffin, E., Harquel, S., Passera, B., Chauvin, A., Bougerol, T., & David, O. (2020). Probing regional cortical excitability via input-output properties using transcranial magnetic stimulation and electroencephalography coupling. *Human Brain Mapping*, *41*(10), 2741‑2761. https://doi.org/10.1002/hbm.24975

Rogasch, N. C., Sullivan, C., Thomson, R. H., Rose, N. S., Bailey, N. W., Fitzgerald, P. B., Farzan, F., & Hernandez-Pavon, J. C. (2017). Analysing concurrent transcranial magnetic stimulation and electroencephalographic data : A review and introduction to the open-source TESA software. *NeuroImage*, *147*, 934‑951. https://doi.org/10.1016/j.neuroimage.2016.10.031

Snyder, D. B., Schmit, B. D., Hyngstrom, A. S., & Beardsley, S. A. (2021). Electroencephalography resting-state networks in people with Stroke. *Brain and Behavior*, *11*(5), e02097. https://doi.org/10.1002/brb3.2097

Tadel, F., Baillet, S., Mosher, J. C., Pantazis, D., & Leahy, R. M. (2011). Brainstorm : A user-friendly application for MEG/EEG analysis. *Computational Intelligence and Neuroscience*, *2011*, 879716. https://doi.org/10.1155/2011/879716

Tangwiriyasakul, C., Premoli, I., Spyrou, L., Chin, R. F., Escudero, J., & Richardson, M. P. (2019). Tensor decomposition of TMS-induced EEG oscillations reveals data-driven profiles of antiepileptic drug effects. *Scientific Reports*, *9*(1), 17057. https://doi.org/10.1038/s41598-019-53565-9

ter Braack, E. M., de Vos, C. C., & van Putten, M. J. A. M. (2015). Masking the Auditory Evoked Potential in TMS-EEG : A Comparison of Various Methods. *Brain Topography*, *28*(3), 520‑528. https://doi.org/10.1007/s10548-013-0312-z

Tremblay, S., Rogasch, N. C., Premoli, I., Blumberger, D. M., Casarotto, S., Chen, R., Di Lazzaro, V., Farzan, F., Ferrarelli, F., Fitzgerald, P. B., Hui, J., Ilmoniemi, R. J., Kimiskidis, V. K., Kugiumtzis, D., Lioumis, P., Pascual-Leone, A., Pellicciari, M. C., Rajji, T., Thut, G., … Daskalakis, Z. J. (2019). Clinical utility and prospective of TMS-EEG. *Clinical Neurophysiology: Official Journal of the International Federation of Clinical Neurophysiology*, *130*(5), 802‑844. https://doi.org/10.1016/j.clinph.2019.01.001

Veniero, D., Bortoletto, M., & Miniussi, C. (2009). TMS-EEG co-registration : On TMS-induced artifact. *Clinical Neurophysiology: Official Journal of the International Federation of Clinical Neurophysiology*, *120*(7), 1392‑1399. https://doi.org/10.1016/j.clinph.2009.04.023
